# Documented Goals of Care Conversations with Hospitalized Patients after Severe Stroke

**DOI:** 10.1101/2023.09.18.23295759

**Authors:** Amber R. Comer, Stephanie Bartlett, Robert Holloway, Claire J. Creutzfeldt, Linda S. Williams, James E. Slaven, Lynn D’Cruz, Katlyn Endris, McKenzie Marchand, Isabel Zepeda, Sumeet Toor, Carly Waite, Areeba Jawed, Alexia M. Torke

## Abstract

**Documented Goals of Care Conversations with Hospitalized Patients after Severe Stroke:** *Background:* Identifying goals of care is important for patients suffering severe ischemic stroke (SIS) and their caregivers to ensure patient- and family-centered treatment decisions. This study sought to determine the prevalence and patient predictors associated with having a documented goals-of-care conversation (dGOCC) after SIS.

*Methods:* We reviewed the medical charts of all patients with National Institutes of Health Stroke Scale (NIHSS) ≥10 admitted to four hospitals in the Midwestern US. In addition to sociodemographic and clinical characteristics, we searched for dGOCC during the acute stroke hospitalization, defined as any documented conversation or meeting that addressed one or more of the following domains: prognostic information, treatment plan, patient preferences and values, quality of life, or establishing goals. We determined prevalence, frequency, timing, and content of dGOCC’s. Additionally, we obtained information on treatment utilization and outcomes.

*Results:* Among 1297 patients, 26.5% (n=344) had at least one dGOCC. Treatment plan was the most discussed domain (n=264, 20% of all patients) and was the most common first dGOCC (n=207, 60% of first conversations). Median day for first dGOCC was on hospital day zero. Patient preferences, values, and goals were documented in 112 (8.6%) of all patients’ charts and quality of life conversations were documented in only 61 (4.7%) charts. In multivariate analysis, having a NIHSS ≥21 (OR 1.46, p-value.01) was associated with having a dGOCC.

*Conclusion:* After severe stroke, most patients do not have a dGOCC, despite the important decisions that often arise about treatment and rehabilitation. Documentation of patient preferences, values and goals are even rarer. This suggests missed opportunities for high quality decision making informed by patient goals to improve person centered care.

## Background

After severe, acute ischemic stroke, patients, families, and clinicians face difficult, time-sensitive decisions about whether to continue or forgo life sustaining treatments (1–2), undergo procedures such as tracheotomy or enteral tube placement, and whether to pursue post-hospital rehabilitation. Most patients who die soon after stroke do so in a hospital with most deaths preceded by a decision to withhold or withdraw life-sustaining treatment (3–5). Setting goals of care is imperative to help patients suffering severe stroke and their caregivers ensure goal-concordant treatment decisions (6–7). Failure to discuss goals of care, or delaying these conversations, may result in a treatment plan that does not align with patient preferences, values, or goals (7–8).

Goals of care conversations (GOCC) contain several components including: 1) effective communication to the patient and family about complex information regarding the stroke and its prognosis; 2) identifying the patient’s individual preferences, values, and goals (“preferences”); 3) providing emotional support to patients and families; 4) engaging in shared medical decision making; and 5) goal setting to ensure treatments and outcomes are aligned with patient and family preferences (6, 9). For GOCCs to be effective, they must explore preferences prior to considering specific treatment interventions. For example, defining a “good” treatment outcome should include exploring an individual patient’s perceptions about their definition of and importance placed on quality of life, because individuals have differing viewpoints on the value of an earlier death vs. survival with poor quality of life (6, 9–10).

One important component of having a GOCC is documenting the conversation (dGOCC) in the patient’s medical record so that the patient’s preferences and goals for medical treatment are known to the entire clinical care team (11). Although the value of GOCCs after stroke is often discussed in the literature, and several challenges to having these conversations have been identified (12–15), only a few studies have examined the documentation of GOCCs in any disease course, (16–17) so little is known about the prevalence and type of dGOCCs after stroke. Additionally, we are unaware of prior studies that have looked at patient factors related to having a dGOCC after stroke. Given the importance of having and documenting GOCCs, this study sought to examine the prevalence and patient characteristics associated with having a dGOCC after severe ischemic stroke (SIS).

## Methods

Adult patients (age ≥18) with a discharge diagnosis of ischemic stroke (International Classification of Diseases-Tenth Revision (ICD-10) inpatient discharge code I-163 and I-164), including patients who survived to discharge and patients who experienced in hospital mortality from four large hospitals between January 1, 2016, and December 31, 2018, who had an initial National Institutes of Health Stroke Scale (NIHSS) score of ≥10 were eligible for this study. We excluded patients with NIHSS below 10 because the need for acute GOCCs in these patients is less clear. We also excluded those without an initial NIHSS documented in the medical record within 24 hours of hospitalization. The four hospitals include 625-bed, 315 bed, 462-bed, and 825 bed centers. Two hospitals have Joint Commission Comprehensive Stroke Center Certifications, and two hospitals have Primary Stroke Center Certifications. Three of the four hospitals were thrombectomy-capable at the time of the study. The Indiana University IRB approved this study.

We used a standardized chart review tool to collect data about clinical severity, including first and worst NIHSS, medical interventions (receipt of tissue plasminogen activator (tPA), thrombectomy, mechanical ventilation, artificial nutrition and hydration), treatment outcomes (length of hospital stay, in hospital mortality, discharge location, including discharge to hospice), and dGOCCs, including content of the dGOCC, parties documented as being present during the conversation, and timing of the conversation during the hospitalization.

The investigator team developed a reproducible method for identifying goals of care conversations from progress notes for chart extraction and classification from admission to diagnosis or death. First, we operationalized goals of care conversations by defining five conversation domains: 1) Treatment Plan; 2) Prognosis; 3) Patient Preferences, Values, and Goals; 4) Patient’s Quality of Life; and 5) Establishing Goals of Medical Care (Figure 1). Next, within each domain, we identified quantifiable examples for each conversation domain called “conversation subtopics” (Figure 1). At least one subtopic within the domain must have been documented as being discussed in order for a conversation to be considered a GOCC. It is possible for a single GOCC to include more than one domain and subtopic. In order for a conversation to qualify as a dGOCC, the note must specify that the clinician had a conversation with or spoke to the patient and/or the patient’s support person. Using the terminology “goals of care conversation” was not required to qualify, as the intention of having a GOCC was inferred from documentation that a conversation or discussion involving one of the five GOCC domains occurred. For example, a dGOCC could begin “spoke with the patient’s wife about the continued use of mechanical ventilation …”. An example of content included in a dGOCC about both preferences and quality of life might include, “while discussing the patient’s inability to care for herself at home if we pursued a PEG tube, the patient’s son told me ‘my mom valued her independence and would not want to live like this.’”

**Figure 1.**
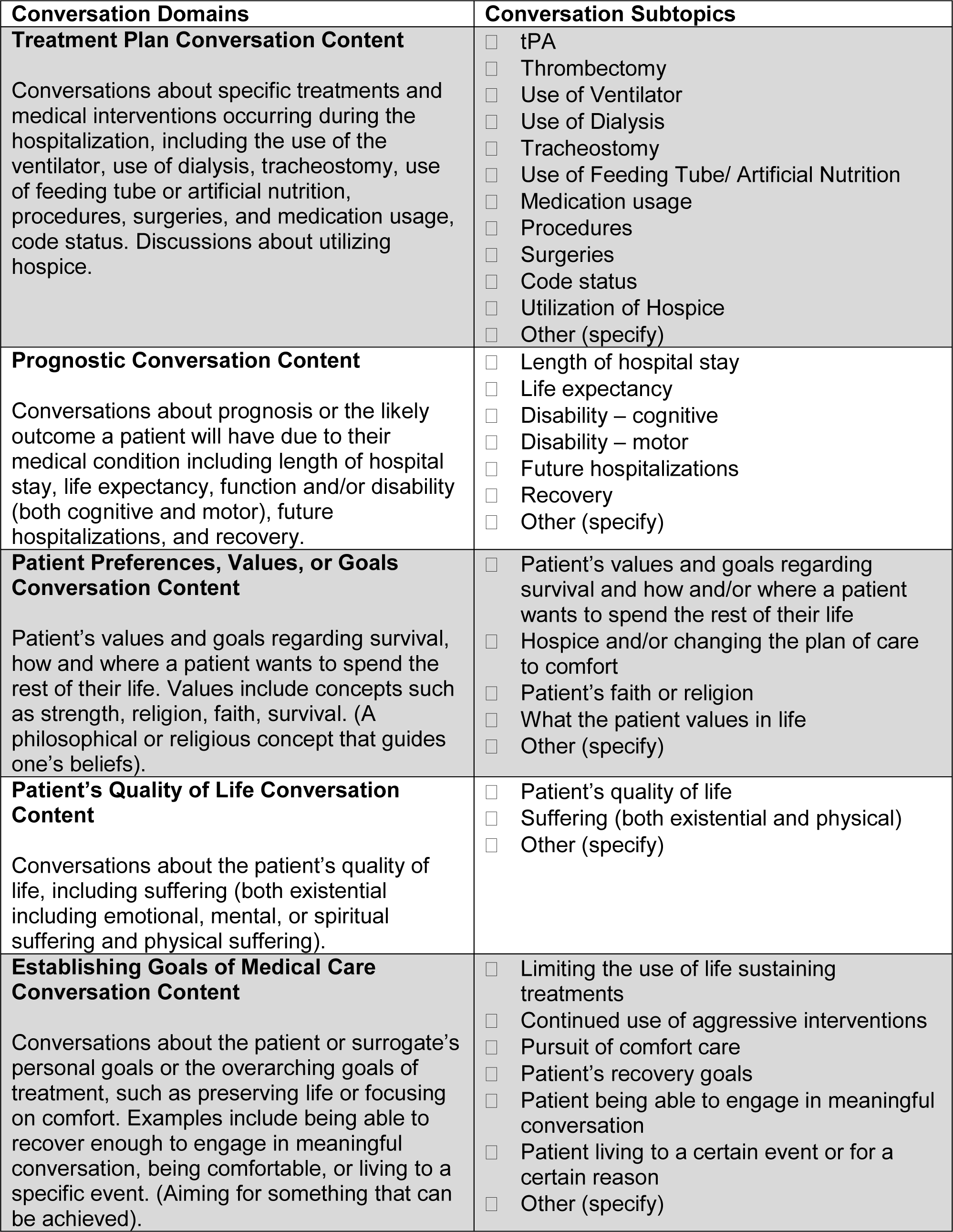
Goals of Care Conversation, Domains and Subtopics

To establish reliability, the PI and all chart reviewers went through manualized training which included learning the structured process for identifying a GOCC. After training, each chart reviewer independently reviewed the same 20 charts. We calculated a kappa statistic on the training charts to ensure interrater reliability was greater than 90% prior to the start of data collection. To ensure fidelity during data collection, charts were randomly checked by the study PI and the chart reviewers met to address questions and ensure continued agreement on dGOCCs. In addition to collecting dGOCCs within the defined domains and subtopics, we documented timing of the conversation during hospitalization, parties documented as being present during the conversation, and physician specialty documenting the conversation.

We calculated descriptive statistics to determine the prevalence of dGOCCs in the patient’s medical records. In bivaratie analysis, we used Chi-Square tests for categorical variables and Student’s t-tests or Wilcoxon rank-sum tests for continuous variables to compare demographic and clinical characteristics between those with and without a documented goals of care conversation. A multivariate logistic regression was performed with dGOCC as the independent variable and sociodemographic and clinical characteristics as the dependent variables. This was performed based on the results of the bivariate associations using logistic regression models, with hospital included as a random effect. Variables were selected for the multivariate analysis both a priori (based on clinician experience) and including those with significance of ≤ .02 in bivariate analysis. NIHSS was dichotomized into a score of 10 – 20, indicating a moderately severe to severe stroke and >=21 indicating a very severe stroke. All analytic assumptions were verified, and all analyses were performed using SAS v9.4 (SAS Institute, Cary, NC).

## Results

A total of 1297 patients were included in this study (Table 1). The majority of participants (54%, n=698) were women, the mean age was 69 years (SD 17), and the median initial NIHSS was 17 (IQR 13-22). More than half of patients experienced their first stroke (61%, n=789), 33% (n=430) of patients received thrombolysis, and 32% (n=405) underwent thrombectomy.

**Table 1.** Bivariate and Multivariate Predictors of Goals of Care Conversations During Hospitalization after.

| Patient Characteristics | All Patients<br>(n=1297) | Patients with<br>GOC<br>(n=344;<br>26.5%) | Patients<br>w/out GOC<br>(n=953;<br>73.5%) | p-value | Multivariate<br>OR (95% CI) | Multivariate<br>p-value |
| --- | --- | --- | --- | --- | --- | --- |
| **Gender** |  |  |  |  |  |  |
| **Women** | 698 (53.9) | 168 (24.1) | 530 (75.9) | .0330* | 0.78 (0.60, 1.01) | .0543 |
| **Men** | 597 (46.1) | 175 (29.3) | 422 (70.7) |  | Reference |  |
| **Age (Mean)** | 68.6 (17.4) | 67.2 (17.1) | 69.1 (17.5) | .0912 | 1.00 (0.99, 1.00) | .1807 |
| **Race** |  |  |  |  |  |  |
| **White** | 1046 (81.2) | 281 (26.9) | 765 (73.1) | .5440 | Reference |  |
| **Black** | 192 (14.9) | 45 (23.4) | 147 (76.6) |  | 0.79 (0.55, 1.15) | .5194 |
| **Other/ Unknown** | 51 (4.0) | 15 (29.4) | 36 (70.6) |  | 0.87 (0.44, 1.71) | .9334 |
| **Initial NIH Stroke Scale**<br>**Score (Median)** | 17 (13, 22) | 18 (13, 23) | 17 (13, 22) | .0959 | ----- | ----- |
| **NIHSS (categorical)** |  |  |  |  |  |  |
| **10 – 20** | 848 (65.4) | 207 (24.4) | 641 (75.6) | .0179* | Reference |  |
| **≥ 21** | 449 (34.6) | 137 (30.5) | 312 (69.5) |  | 1.46 (1.11, 1.91) | **.0065\*** |
| **Prior CVA/TIA** |  |  |  |  |  |  |
| **First Stroke (no prior)** | 789 (61.2) | 221 (28.0) | 568 (72.0) | .1072 | Reference |  |
| **Secondary Stroke (yesPrior)** | 501 (38.8) | 120 (24.0) | 381 (76.0) |  | 0.84 (0.64, 1.10) | .2048 |
| **tPA** |  |  |  |  |  |  |
| **Yes** | 430 (33.3) | 124 (28.8) | 306 (71.2) | .1997 | 1.03 (0.77, 1.37) | .8533 |
| **No** | 863 (66.7) | 220 (25.5) | 643 (74.5) |  | Reference |  |
| **Thrombectomy** |  |  |  |  |  |  |
| **Yes** | 405 (31.6) | 118 (29.1) | 287 (70.9) | .1463 | 0.96 (0.71, 1.29) | .7773 |
| **No** | 878 (68.4) | 222 (25.3) | 656 (74.7) |  | Reference |  |
| **Hospital Location** |  |  |  |  |  |  |
| **Hospital 1** | 106 (8.2) | 26 (24.5) | 80 (75.5) | .004* |  |  |
| **Hospital 2** | 210 (16.2) | 43 (20.5) | 167 (79.5) |  |  |  |
| **Hospital 3** | 563 (43.4) | 139 (24.7) | 424 (75.3) |  |  |  |
| **Hospital 4** | 418 (32.2) | 136 (32.5) | 282 (67.5) |  |  |  |

### Documented Goals of Care Conversations

Among all patients, 26.5% (n=344), had a goals of care conversation documented in their medical record (Table 1). Among those with a dGOCC, 39% (n=135) had only one conversation documented (Figure 2). The majority of first dGOCCs (75%) were held on or before hospital day 7; however, one out of four first GOCCs (25%) occurred on or after hospital day 8. The majority of dGOCCs were documented by non-clinician members of the care team, including chaplains and social workers. Among physician specialties, internal medicine had the highest prevalence of dGOCCs (52%, n=152) (Table 3). Adult children were the most common support person documented as being present during the conversation (documented as being present in 41% of GOCCs) (Table 3).

**Figure 2.**
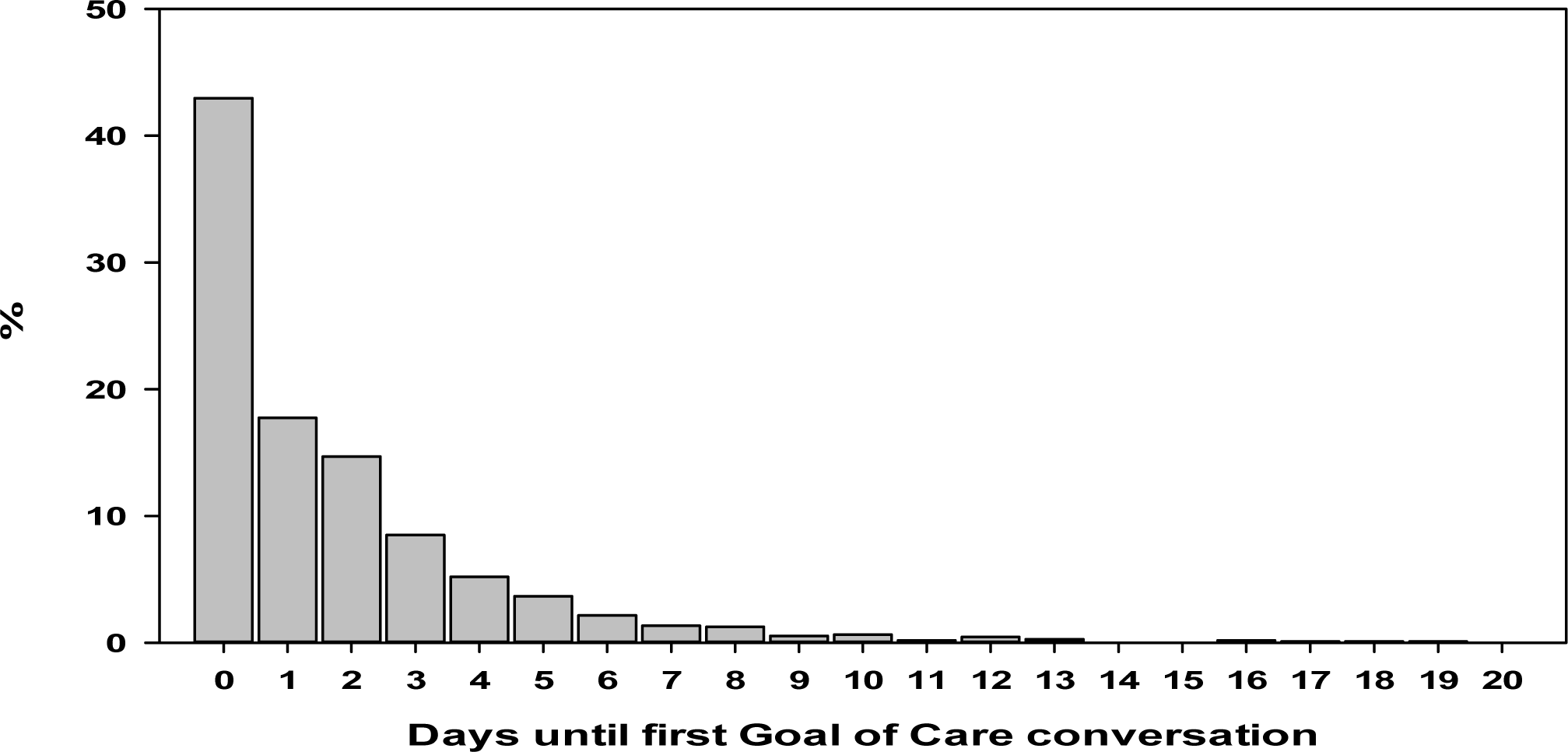
GOCC after Severe Stroke

### Demographic and Clinical Factors Associated with a dGOCC

In bivariate analysis, compared to those without dGOCC, men (p-value = .03) and those with a high NIHSS (p-value=.02) were more likely to have a dGOCC. We also observed significant variation in the proportion of SIS patients with a dGOCC between hospitals (p-value=<.01) (Table 1). In multivariate analysis, higher NIHSS was the only variable associated with having a dGOCC (Adjusted Odds Ratio 1.46, 95% Confidence Interval 1.11,1.91, p<.01) (Table 1).

### Goals of Care Conversation Content

Among patients with a dGOCC, treatment plan was the most common dGOCC domain discussed, occurring in 77% (n=264) of patients who had a dGOCC (Table 2 and Figure 3). Establishing medical treatment goals was the second most common domain discussed during dGOCC (n=233, 68%). Patient preferences (33%, n=112) and Quality of life (18%, n=61) were the domains discussed the least (Table 2 and Figure 3). Among all severe stroke patients, the patient’s quality of life (including conversations about past, current, or future quality of life) was only documented as being discussed with 4.7% (n=61) of patients; and patients’ preferences were documented as being discussed with only 8.6% of all severe stroke patients (n=112) (Table 2). Hospital location was significantly associated with dGOCC domains including treatment plan (p-value>.01), patient preferences (p-value>.01), and establishing goals (p-value>.01).

**Table 2.** Patient Characteristics and Bivariate Analysis of Characteristics Associated with GOCC Domains.

|  | Prognostic Conversation<br>(n=126; 9.7%) |  |  | Patients with Treatment<br>Plan Conversation<br>(n=264; 20.3%) |  |  | Patients with Quality-of-Life Conversation<br>(n=61; 4.7%) |  |  | Patients with Patient<br>Preferences, Values, and<br>Goals Conversation<br>(n=112; 8.6%) |  |  | Patients with Establishing<br>Goals Conversation<br>(n=233; 18.0%) |  |  |
| --- | --- | --- | --- | --- | --- | --- | --- | --- | --- | --- | --- | --- | --- | --- | --- |
| Patient<br>Characteristics | Yes | No | p-value | Yes | No | p-value | Yes | No | p-value | Yes | No | p-value | Yes | No | p-value |
| Gender |  |  |  |  |  |  |  |  |  |  |  |  |  |  |  |
| Women | 52<br>(7.5) | 646<br>(92.6) | .0037* | 125<br>(17.9) | 573<br>(82.1) | .0202* | 31<br>(4.4) | 667<br>(95.6) | .7224 | 52<br>(7.4) | 646<br>(92.6) | .1190 | 117<br>(16.8) | 581<br>(83.2) | .2421 |
| Men | 73<br>(12.2) | 524<br>(87.8) |  | 138<br>(23.1) | 459<br>(76.9) |  | 29<br>(4.9) | 568<br>(95.1) |  | 59<br>(9.9) | 538<br>(90.1) |  | 115<br>(19.3) | 482<br>(80.7) |  |
| Age (Mean) | 64.3<br>(18.4) | 69.0<br>(17.3) | .0043* | 67.9<br>(68.7) | 68.7<br>(17.5) | .4799 | 69.3<br>(15.3) | 68.5<br>(17.5) | .7462 | 68.1<br>(16.5) | 68.6<br>(17.5) | .7609 | 68.0<br>(18.0) | 68.7<br>(17.3) | .5612 |
| Race |  |  |  |  |  |  |  |  |  |  |  |  |  |  |  |
| White | 99<br>(9.5) | 947<br>(90.5) | .6010 | 215<br>(20.5) | 831<br>(79.5) | .4304 | 48<br>(4.6) | 998<br>(95.4) | .1740 | 91<br>(8.7) | 955<br>(91.3) | .6723 | 196<br>(18.7) | 850<br>(81.3) | .0920 |
| Black | 19<br>(9.9) | 173<br>(90.1) |  | 34<br>(17.7) | 158<br>(82.3) |  | 7<br>(3.6) | 185<br>(96.4) |  | 15<br>(7.8) | 177<br>(92.2) |  | 24<br>(12.5) | 168<br>(87.5) |  |
| Other/<br>Unknown | 7<br>(13.7) | 44<br>(86.3) |  | 13<br>(25.5) | 38<br>(74.5) |  | 5<br>(9.8) | 46<br>(90.2) |  | 6<br>(11.8) | 45<br>(88.2) |  | 11<br>(21.6) | 40<br>(78.4) |  |
| Initial NIH<br>Stroke Scale<br>Score (Median) | 21.5<br>(15, 25) | 17<br>(13, 22) | <.0001* | 18<br>(13, 24) | 17<br>(13, 22) | .0047* | 22<br>(15, 26) | 17<br>(13, 22) | .0010* | 20<br>(13, 25) | 17<br>(13, 22) | .0017* | 19<br>(14, 24) | 17<br>(12.5, 22) | .0006* |
| NIHSS<br>(categorical) |  |  |  |  |  |  |  |  |  |  |  |  |  |  |  |
| 10 – 20 | 59<br>(7.0) | 789<br>(93.0) | <.0001* | 149<br>(17.6) | 699<br>(82.4) | .0006* | 26<br>(3.1) | 822<br>(96.9) | .0001* | 57<br>(6.7) | 791<br>(98.3) | .0007* | 129<br>(15.2) | 719<br>(84.8) | .0004* |
| >=21 | 67<br>(14.9) | 382<br>(85.1) |  | 115<br>(25.6) | 334<br>(74.4) |  | 35<br>(7.8) | 414<br>(92.2) |  | 55<br>(12.2) | 394<br>(87.8) |  | 104<br>(23.2) | 345<br>(76.8) |  |
| Prior CVA/TIA |  |  |  |  |  |  |  |  |  |  |  |  |  |  |  |
| First Stroke -<br>NO | 90<br>(11.4) | 699<br>(88.6) | .0128* | 164<br>(20.8) | 625<br>(79.2) | .5940 | 38<br>(4.8) | 751<br>(95.2) | .8525 | 77<br>(9.8) | 712<br>(90.2) | .0635 | 149<br>(18.9) | 640<br>(81.1) | .2140 |
| First Stroke -<br>YES | 36<br>(7.2) | 465<br>(92.8) |  | 98<br>(19.6) | 403<br>(80.4) |  | 23<br>(4.6) | 478<br>(95.4) |  | 34<br>(6.8) | 467<br>(93.2) |  | 81<br>(16.2) | 420<br>(83.8) |  |
| tPA | 37<br>(8.6) | 393<br>(91.4) | .3292 | 101<br>(23.5) | 329<br>(76.5) | .0532 | 14<br>(3.3) | 416<br>(96.7) | .0801 | 42<br>(9.8) | 388<br>(90.2) | .3185 | 73<br>(17.0) | 357<br>(83.0) | .4908 |
| Thrombectomy | 49<br>(12.1) | 356<br>(87.9) | .0379* | 91<br>(22.5) | 314<br>(77.5) | .1822 | 19<br>(4.7) | 386<br>(95.3) | .9864 | 44<br>(10.9) | 361<br>(89.1) | .0555 | 78<br>(19.3) | 327<br>(80.7) | .3702 |
| Hospital<br>Location |  |  |  |  |  |  |  |  |  |  |  |  |  |  |  |
| Hospital 1 | 11<br>(10.4) | 95<br>(89.6) | .0212* | 23<br>(21.7) | 83<br>(78.3) | .7235 | 7<br>(6.6) | 99<br>(93.4) | .7350 | 13<br>(12.3) | 93<br>(87.7) | <.0001* | 19<br>(17.9) | 87<br>(82.1) | <.0001* |
| Hospital 2 | 9<br>(4.3) | 201<br>(95.7) |  | 42<br>(20.0) | 168<br>(80.0) |  | 9<br>(4.3) | 201<br>(95.7) |  | 18<br>(8.6) | 192<br>(91.4) |  | 22<br>(10.5) | 188<br>(89.5) |  |
| Hospital 3 | 66<br>(11.7) | 497<br>(88.3) |  | 121<br>(21.5) | 442<br>(78.5) |  | 24<br>(4.3) | 539<br>(95.7) |  | 71<br>(12.6) | 492<br>(87.4) |  | 71<br>(12.6) | 492<br>(87.4) |  |
| Hospital 4 | 40<br>(9.6) | 378<br>(90.4) |  | 78<br>(18.7) | 340<br>(81.3) |  | 21<br>(5.0) | 397<br>(95.0) |  | 10<br>(2.4) | 408<br>(97.6) |  | 121<br>(28.9) | 297<br>(71.1) |  |

**Table 3.**
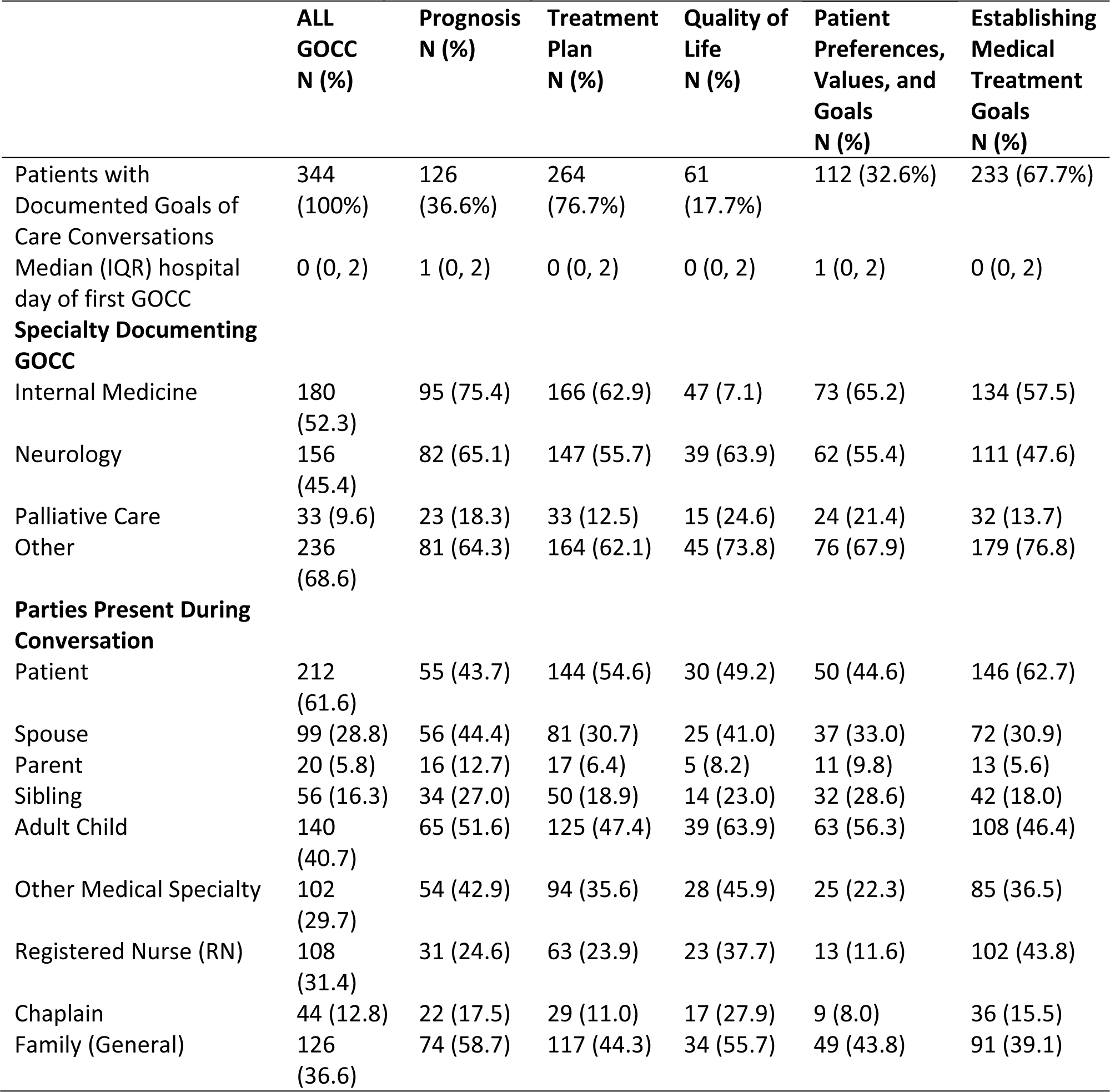
Domains and Specialty conducting Goals of Care Conversations.

|  | **ALL**<br>**GOCC**<br>**N (%)** | **Prognosis**<br>**N (%)** | **Treatment**<br>**Plan**<br>**N (%)** | **Quality of**<br>**Life**<br>**N (%)** | **Patient**<br>**Preferences,**<br>**Values, and**<br>**Goals**<br>**N (%)** | **Establishing**<br>**Medical**<br>**Treatment**<br>**Goals**<br>**N (%)** |
| --- | --- | --- | --- | --- | --- | --- |
| Patients with Documented Goals of Care Conversations | 344<br>(100%) | 126<br>(36.6%) | 264<br>(76.7%) | 61<br>(17.7%) | 112 (32.6%) | 233 (67.7%) |
| Median (IQR) hospital day of first GOCC | 0 (0, 2) | 1 (0, 2) | 0 (0, 2) | 0 (0, 2) | 1 (0, 2) | 0 (0, 2) |
| **Specialty Documenting GOCC** |  |  |  |  |  |  |
| Internal Medicine | 180<br>(52.3) | 95 (75.4) | 166 (62.9) | 47 (7.1) | 73 (65.2) | 134 (57.5) |
| Neurology | 156<br>(45.4) | 82 (65.1) | 147 (55.7) | 39 (63.9) | 62 (55.4) | 111 (47.6) |
| Palliative Care | 33 (9.6) | 23 (18.3) | 33 (12.5) | 15 (24.6) | 24 (21.4) | 32 (13.7) |
| Other | 236<br>(68.6) | 81 (64.3) | 164 (62.1) | 45 (73.8) | 76 (67.9) | 179 (76.8) |
| **Parties Present During Conversation** |  |  |  |  |  |  |
| Patient | 212<br>(61.6) | 55 (43.7) | 144 (54.6) | 30 (49.2) | 50 (44.6) | 146 (62.7) |
| Spouse | 99 (28.8) | 56 (44.4) | 81 (30.7) | 25 (41.0) | 37 (33.0) | 72 (30.9) |
| Parent | 20 (5.8) | 16 (12.7) | 17 (6.4) | 5 (8.2) | 11 (9.8) | 13 (5.6) |
| Sibling | 56 (16.3) | 34 (27.0) | 50 (18.9) | 14 (23.0) | 32 (28.6) | 42 (18.0) |
| Adult Child | 140<br>(40.7) | 65 (51.6) | 125 (47.4) | 39 (63.9) | 63 (56.3) | 108 (46.4) |
| Other Medical Specialty | 102<br>(29.7) | 54 (42.9) | 94 (35.6) | 28 (45.9) | 25 (22.3) | 85 (36.5) |
| Registered Nurse (RN) | 108<br>(31.4) | 31 (24.6) | 63 (23.9) | 23 (37.7) | 13 (11.6) | 102 (43.8) |
| Chaplain | 44 (12.8) | 22 (17.5) | 29 (11.0) | 17 (27.9) | 9 (8.0) | 36 (15.5) |
| Family (General) | 126<br>(36.6) | 74 (58.7) | 117 (44.3) | 34 (55.7) | 49 (43.8) | 91 (39.1) |

**Figure 3.**
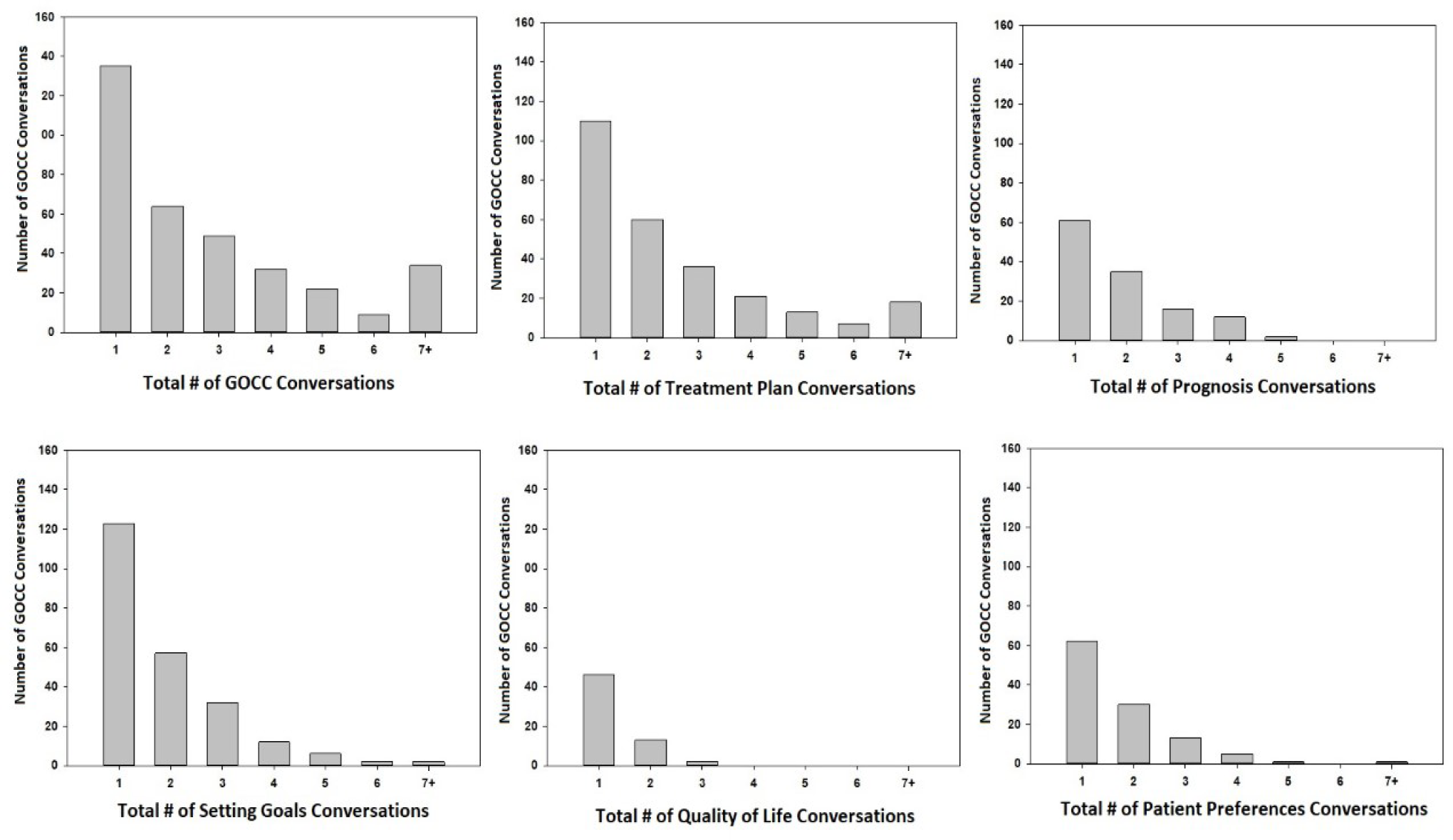
Number of GOCC conversations and GOCC content

## Discussion

Our study found a low prevalence of dGOCC after severe stroke. The results of this study are similar to studies that have identified a low prevalence of documented goals of care conversations in other life-threatening conditions (16–19). This finding has two possible causes: GOCC are occurring but not being documented (or identified), or GOCC are not occurring at high rates. Based on prior studies, both causes are likely to be at play. Prior studies have found both physicians and patients report a low rate of GOCC in critically ill patients, and prior studies report a low prevalence of GOCC documented in the medical record (19). Among the few patients with a GOCC, most patients only had one conversation and the conversation tended to be early during the hospitalization. In this study, the only predictor associated with having a dGOCC was high NIHSS. It is appropriate that most dGOCC are occurring early and after the most severe strokes, but there are likely missed opportunities for all patients, including those who have lower NIHSS but nonetheless have disabling symptoms or are facing major medical decisions (20).

This study had several limitations. First, this study was only able to account for dGOCC. Therefore, the low number of conversations reported may be due to documentation failure and not due to the conversations not occurring. Although it is possible that the number of GOCC may be higher, a focus on documented conversations is important because critical information about patient preferences that is not documented may not be transmitted to other members of the patient’s clinical care team, leading to care that is not congruent with patient preferences. Although we conducted the study in multiple hospitals with different providers and systems of care, all were in one Midwestern city and the population included a small number of non-white patients, thus the findings may not be generalizable to other areas of the United States or other hospital systems of care. Lastly, we developed and applied a structured definition of GOCC, but it is possible that conversations which others may consider to be part of GOCC were not categorized and recorded as such in this study. We attempted to minimize this possibility by intentionally using a broad and inclusive definition of GOCC. Future studies are needed to determine best practices in conducting and documenting GOCC and to further study patient-centered outcomes, including goal concordant care, associated with having dGOCC during hospitalization for SIS.

This study of almost 1300 patients with SIS found a dGOCC in the medical record of only 26.5% of patients. Given that SIS is a major life-altering event that leads to changes in independent living, quality of life and cognition, these findings suggest there are likely missed opportunities for having GOCC with this cohort of patients and/or their surrogate medical decision makers. Although we may have missed conversations that occurred but were not documented, implementing interventions to increase both the frequency and the documentation of GOCC may be beneficial. Taking steps to improve GOCC frequency and documentation is important because dGOCCs will likely aid communication between multiple teams caring for patients with SIS and are associated with reduced ICU utilization and increased goal concordant care (21).

## Data Availability

Data can be made available by contacting the corresponding author.

## Acknowledgments

None

